# Fundamental Bound on Epidemic Overshoot in the SIR Model

**DOI:** 10.1101/2023.06.02.23290891

**Authors:** Maximilian M. Nguyen, Ari S. Freedman, Sinan A. Ozbay, Simon A. Levin

## Abstract

We derive an exact upper bound on the epidemic overshoot for the Kermack-McKendrick SIR model. This maximal overshoot value of 0.2984… occurs at 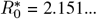. Using the general analysis framework presented within, we then consider more complex SIR models, such as those that incorporate vaccination or contact heterogeneity. We analyze models that consider vaccinations and show that the presence of vaccinated individuals decreases the maximum possible overshoot. For epidemics where the contact structure is given by a network, we numerically find that increased contact heterogeneity lowers the maximal overshoot value and weakens the dependency of overshoot on transmission.

## Introduction

The overshoot of an epidemic is the proportion of the population that becomes infected after the peak of the epidemic has already passed. Formally, it is given as the difference between the fraction of the population that is susceptible at the peak of infection prevalence and at the end of the epidemic. Intuitively, it is the difference between the herd immunity threshold and the total fraction of the population that gets infected [7, 4]. As it describes the damage to the population in the declining phase of the epidemic (i.e. when the effective reproduction number is less than 1), one might be tempted to dismiss its relative importance. However, a substantial proportion of the epidemic, and thus a large number of people, may be impacted during this phase of the epidemic dynamics.

A natural question to ask then is how large can the overshoot be and how does the overshoot depend on epidemic parameters, such as transmissibility and recovery rate? Surprisingly, within the framework of the SIR ODE model, this question can be answered exactly. In this paper, we first derive the bound on the overshoot in the Kermack-McKendrick limit of the SIR model [8]. Beyond the basic SIR model, we then see if the bound on overshoot holds if we add additional complexity, such as vaccinations, into the model. We then numerically study what happens when we move to a more realistic structural model of epidemics that incorporates contact heterogeneity.

## Results

The classical Kermack-McKendrick SIR model is given by the following set of ODEs:

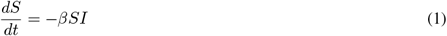

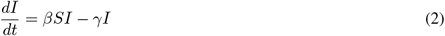

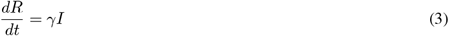

where *S, I*, and *R* are the fractions of population in the susceptible, infected, or recovered state respectively. As these are the only possible states within this model, the conservation equation for the whole population is given as *S* + *I* + *R* = 1. Conceptually, the overshoot can be equivalently calculated in two ways. In the first it is given by the difference in the fraction of susceptible individuals at the peak of infection prevalence 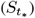 and at the end of the end of the epidemic (*S*_*∞*_) (Figure 1a). Alternatively, it can be viewed as the integration of the number of newly infected individuals, which is given by the infection incidence curve 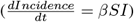 from the peak of infection prevalence to the end of the epidemic (Figure 1b). We will make use of the former relationship in the results that follow.

**Figure 1:**
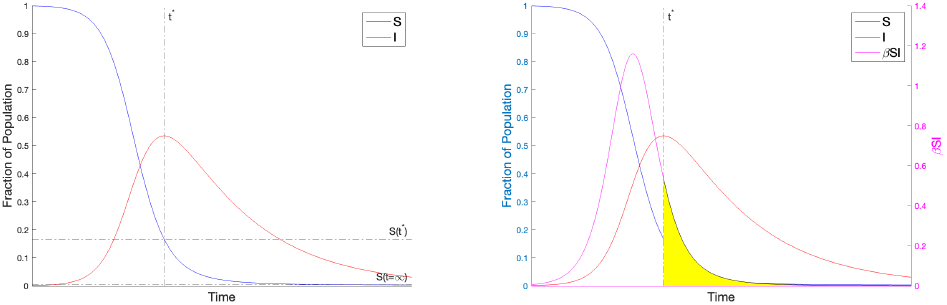
The overshoot can be calculated in two ways: a) Overshoot is calculated as the difference between the fraction of the population that is susceptible at *t*^*\**^ and infinite time. b) Overshoot is calculated as the integral of the infection incidence curve from *t*^*\**^ until infinite time. Therefore overshoot corresponds to the area of the region shaded in yellow.

The only two parameters of this model are *β* and *γ*. A key parameter in epidemic modeling combines these two into a single parameter by taking their ratio, which is known as the basic reproduction number (*R*_0_). While overshoot is a function of both *β* and *γ* independently, the behavior of the overshoot can be parameterized in terms of the single parameter *R*_0_ (Figure S1). Plotting the dependency of overshoot on *R*_0_ (Figure 2), we observe a peak in the curve at 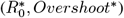 that sets an upper bound on the overshoot. From a public health perspective, diseases that have estimated *R*_0_’s near this peak region in Figure 2 include COVID-19 (ancestral strain) [2], SARS [16], diphtheria [14], monkeypox [6], and ebola [15]. This peak phenomena in the overshoot was first numerically observed by [17], though not explained. We will now derive the solution for this maximum point analytically.

**Figure 2:**
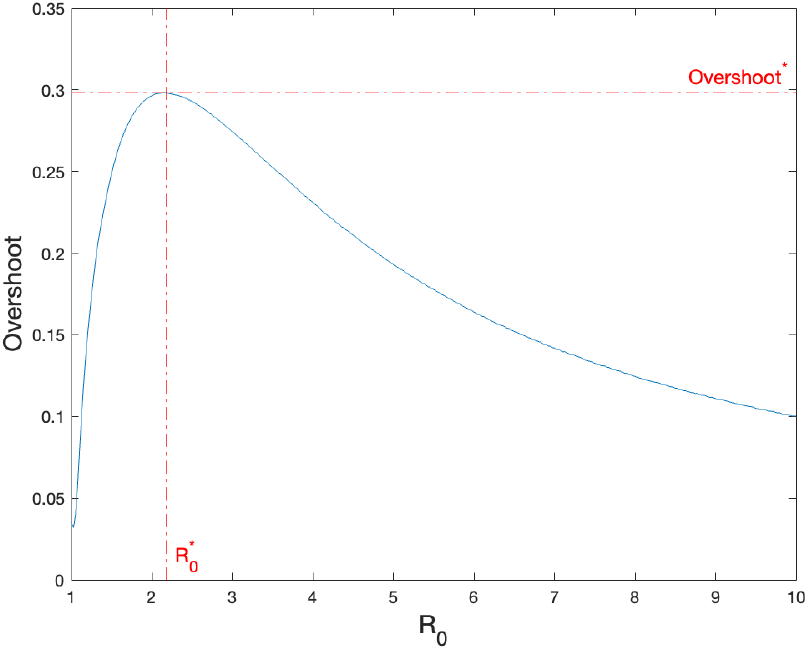
The overshoot as a function of *R*_0_ for the Kermack-McKendrick SIR model.

### Deriving the Exact Bound on Overshoot in the Kermack-McKendrick SIR Model

*Theorem: The maximum possible overshoot in the Kermack-McKendrick SIR model is a fraction* 0.2984… *of the entire population, with a corresponding* 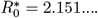.

Proof.

Let *t*^*\**^ be the time at the peak of the infection prevalence curve. The herd immunity threshold is the difference in the fractions of the population that are susceptible at zero time and at *t*^*\**^. Define overshoot as the difference in the fractions of the population that are susceptible at *t*^*\**^ and at infinite time. This is equivalent to defining overshoot as the cumulative fraction of the population that gets infected after *t*^*\**^.

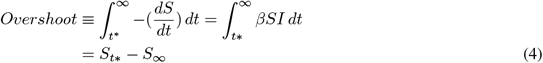

where *S*_*t\**_ and *S*_*∞*_ are the susceptible fractions at *t*^*\**^ and infinite time respectively. We will use 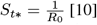 [10], which can be obtained by setting (2) to zero and solving for that critical *S*. We will use the notation *X*_*t*_ to indicate the value of compartment X at time *t*.

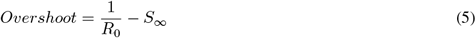

Since we would like to compute maximal overshoot, we can differentiate the overshoot equation (5) with respect to *S*_*∞*_ to find the extremum. We will eliminate *R*_0_ from the overshoot equation so that we have an equation only in terms of *S*_*∞*_.

To find an expression for *R*_0_, we start by deriving the standard final size relation for the SIR model [1, 3]. We solve for the rate of change of I as a function of S using (1)-(2) to obtain

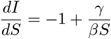

from which it follows on integration that 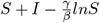 is constant along any trajectory.

Considering the beginning of the epidemic and the peak of the epidemic yields:

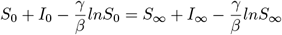

hence

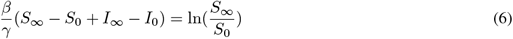

We now define the initial conditions: *S*_0_ = 1 − *ϵ* and *I*_0_ = *ϵ*, where *ϵ* is the (infinitesimally small) fraction of initially infected individuals. We use the standard asymptotic of the SIR model that there are no infected individuals at the end of an SIR epidemic: *I*_*∞*_ = 0. Hence, recalling that 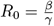 we obtain that

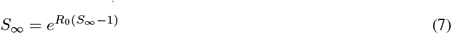

The resulting equation (7) is the final size relation for the Kermack-McKendrick SIR model.

Rearranging for *R*_0_ yields the following expression:

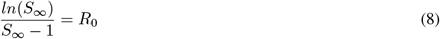

We then substitute this *R*_0_ expression (8) into the overshoot equation (5).

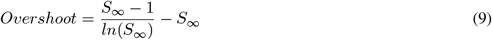

Differentiating with respect to *S*_*∞*_ and setting the equation to zero to find the maximum overshoot yields:

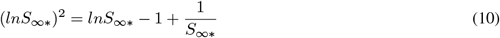

whose solution is

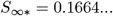

and which corresponds to

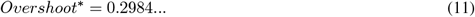

using (9). The corresponding *R*_0_ calculated using (8) is

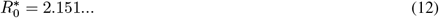

This concludes the proof ◼

Additionally, to find the total recovered fraction is straightforward. In the asymptotic limit of the SIR model, there are no remaining infected individuals, so *R*_*∞\**_ = 1 − *S*_*∞\**_.

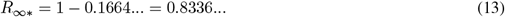

In other words, approximately 5 out of every 6 individuals in the population will have experience infection when overshoot is maximized.

### Upper Bounds on Overshoot in Models that Include Vaccinations

Beyond the Kermack-McKendrick SIR model, one can ask if the bound on overshoot still holds if other complexities are added to the model. First, we will consider the addition of vaccinations.

We will consider three qualitatively different types of curves for the vaccination rate (Figure 3). These correspond to different scenarios that might be modeled. The first model assumes a vaccination rate of zero after the outbreak begins, which implies all vaccinations occur before the outbreak. The second model of vaccination assumes a constant per-capita vaccination rate. This is a situation where all susceptible individuals get vaccinated at the same rate. This assumption yields a vaccination curve for the population that is concave down. The third type of model assumes a risk-driven vaccination rate that depends on the number of infected individuals. This yields a non-monotonic vaccination curve for the population that switches from being initially concave up to being concave down. Depending on the scenario being analyzed, one model might be more appropriate to use than others. Below we discuss each model in further detail by providing the corresponding system of equations, relevant scenarios the model might correspond to in reality, and the corresponding maximal overshoot for each model.

**Figure 3:**
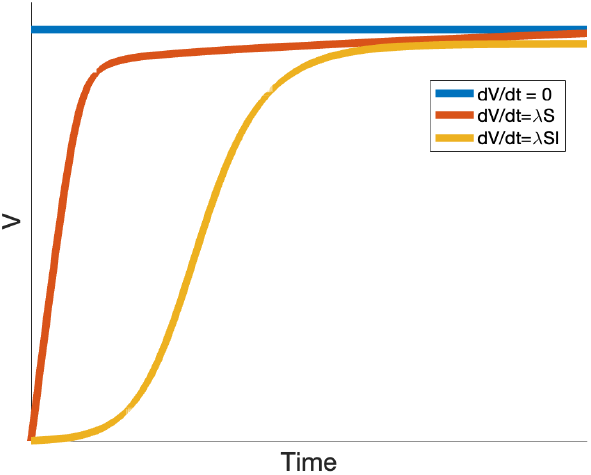
The fraction of population that is vaccinated (V) based on different vaccination rates: a vaccination rate of zero over the course of the epidemic (blue), a constant per-capita vaccination rate (red), and a risk-driven vaccination rate (yellow).

#### Maximal Overshoot when the Number of Vaccinated Individuals is Constant

The first model of vaccination assumes there are no vaccinations during the outbreak, which implies a fixed number of vaccinated individuals over the course of the epidemic. Such a scenario might be the reintroduction of an infectious disease into a population that has a pre-existing level of immunity.

Since the number of vaccinated individuals is constant, this implies all vaccinations occurred prior to the initial time step. The calculation is then trivial assuming vaccinations provide complete and permanent immunity. In that case, vaccinated individuals can simply be ignored entirely in the dynamics, resulting in the maximal overshoot simply scaling with the unvaccinated fraction.

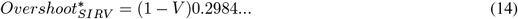

#### Maximal Overshoot Under Addition of Constant Per-Capita Vaccination

We next consider a more typical scenario where the vaccination rate per unvaccinated individual is constant per unit time. Barring any additional information about the population or the epidemic, it is reasonable to assume that all susceptible individuals are vaccinated at the same rate. Consider the following SIRV model:

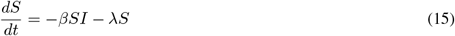

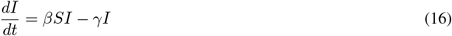

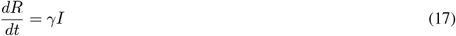

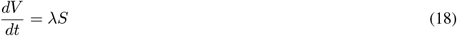

In this case, it is easily shown that there is a conserved quantity, 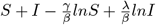, which reduces to (6) when the vaccination rate is zero (i.e. *λ* = 0). Unfortunately, having the conserved quantity is not sufficient to compute the overshoot, since there does not appear to be a way to separate infected and vaccinated individuals when trying to extend the previous calculation. Therefore, we turn to numerical computation (Figure 4a). We find that the maximal overshoot is bounded above by the value already obtained in the model without vaccinations. As shown in Figure 4, the overshoot has a complicated dependence on the vaccination parameter *λ* and *R*_0_.

**Figure 4:**
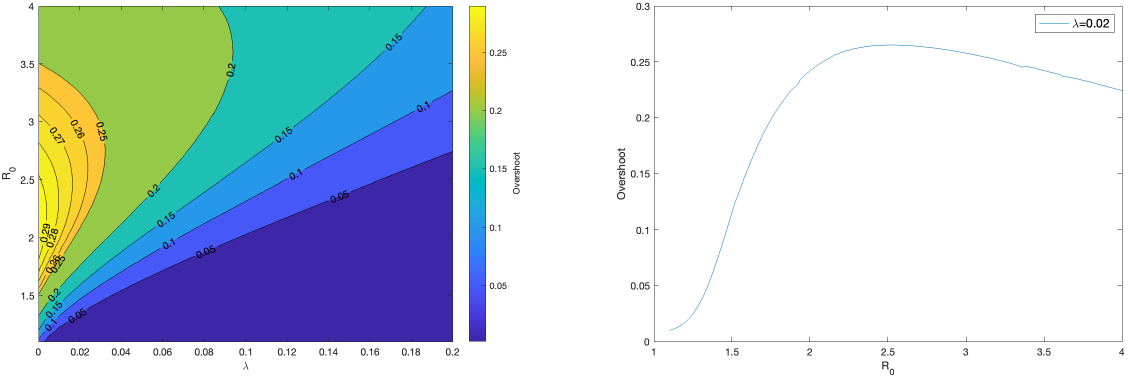
a) Contour plot for the overshoot for the SIRV model with 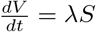 as a function of *λ* and *R*_0_. b) Vertical cross-section of the contour plot from (a) for *λ* = 0.02.

#### Maximal Overshoot Under Addition of a Risk-Driven Vaccination Rate

Lastly consider a vaccination rate that is proportional to the number of infected individuals. Such risk-driven behavior may arise for a variety of reasons, including initial vaccine hesitancy, a delay in vaccine availability, or a correlation between willingness to get vaccinated and the number of infected individuals. Consider the following SIRV model:

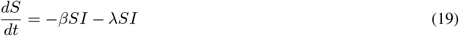

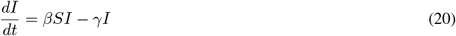

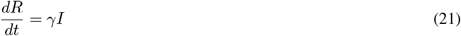

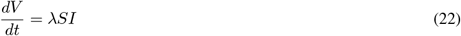

Since the model now has an additional compartment, V, compared with the original SIR model, we must update our definition for overshoot accordingly. Fundamentally, overshoot compares the fraction of people who have not been infected at the epidemic peak and the people who have not been infected at the end of the epidemic. The fraction of people who have not been infected at any particular time, *t*, is *S*_*t*_ + *V*_*t*_. Thus, overshoot can be redefined as follows.

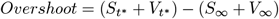

Since the equation for 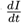 remains unchanged, 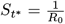 still applies. Thus, the overshoot equation for models with vaccinated compartments is given by:

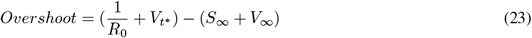

To maximize overshoot, we thus need to find expressions for *R*_0_, 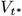, and *V*_*∞*_ in terms of *S*_*∞*_.

To find *R*_0_ we start by taking the ratio 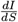 and integrating as before. It follows that 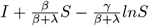 is constant along any trajectory. Considering the beginning and the end of the epidemic yields:

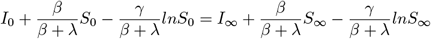

Using the same initial conditions, asymptotic behavior, and parameter substitution as before 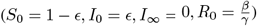 yields the following final size relation.

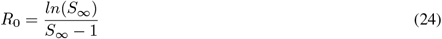

Thus, we see that *R*_0_ for this SIRV model takes on the same expression as the SIR model (8).

To find 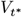, let us take the ratio of time derivatives of the S and V compartments (19), (22),

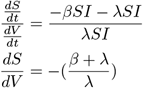

from which it follows on integration that 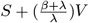 is constant along any trajectory. Considering the beginning and the peak of the epidemic yields:

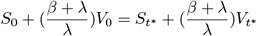

Using the initial conditions (*S*_0_ = 1 − *ϵ, I*_0_ = *ϵ, V*_0_ = 0) and recalling that 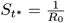, we obtain the following formula for 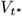.

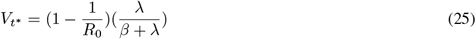

To find *V*_*∞*_, recall that 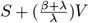 is constant along any trajectory. Considering the peak of the epidemic and the end of the epidemic yields

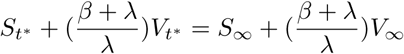

Using the equation for 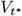 (25) and recalling that 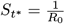, we obtain the following equation for *V*_*∞*_.

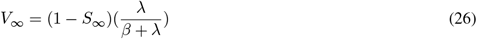

Substituting the expressions for *R*_0_ (24), 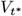 (25), *V*_*∞*_ (26) into the overshoot equation (23) yields:

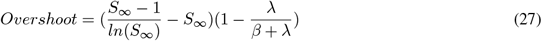

We see that this expression for the overshoot is simply the overshoot expression for the original SIR model (9) scaled by a factor 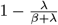.

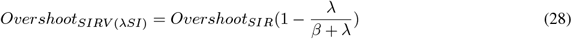

Since both *β, λ* ≥ 0, then the factor 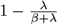 can never be greater than 1. This implies that the bound on maximal overshoot given by the theorem holds, becoming exact in the limit of no vaccinations (ie. *λ* = 0). For this model, the maximal overshoot decreases as a function of *λ* in a nonlinear way and has a nonlinear dependence on *R*_0_ (Figure 5).

**Figure 5:**
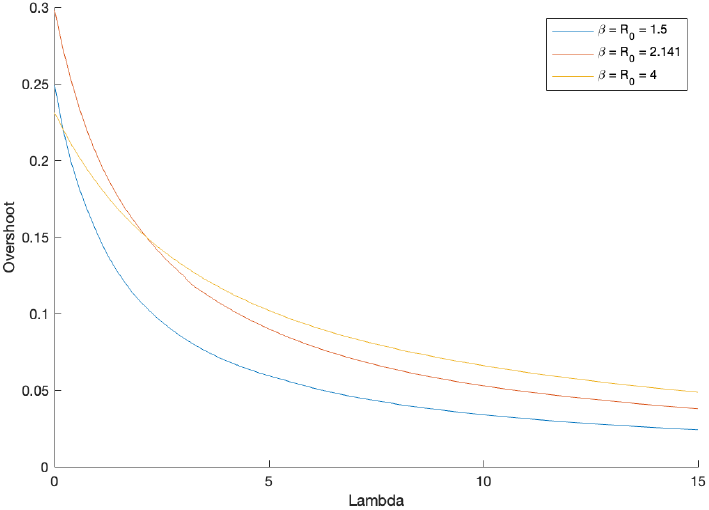
The overshoot for the SIRV model with 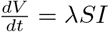 as a function of *λ* for different levels of *β* (or equivalently *R*_0_).

### Overshoot Behavior Under Addition of Contact Heterogeneity

One of the key assumptions of the Kermack-McKendrick model is a well-mixed population where every infected individual has the same effect on the population. While a mathematically convenient assumption, real-world epidemics happen in more structured populations [13]. This is evidenced by the phenomena of superspreaders, where some fraction of infected individuals infect a disproportionately large number of susceptibles. This is modeled in the literature through the introduction of contact heterogeneity [9, 5].

Here we explore what happens to the overshoot when the contact structure of the population is given by a network graph that is roughly one giant component. While it is possible to construct pathological graphs that produce very complex dynamics, we consider more classical graphs here. Using a parameterization of heterogeneity given by Ozbay et. al. [12] (see Methods for details), we simulated epidemics on networks with structure ranging from the homogeneous limit (well-mixed, complete graph) to a heterogeneous limit (heavy-tailed degree distributions). We observed what happens to the overshoot on these different graphs as we changed the transmission probability.

On a network model, where contact structure is made explicit, the homogeneous graph is a complete graph, which recapitulates the well-mixed assumption of the Kermack-McKendrick model. It is not surprising then that the overshoot in the homogeneous graph (*σ* = 0) peaks also around 0.3 (Figure 6, *green*) and has qualitatively the same shape profile as the ODE model (Figure 2).

**Figure 6:**
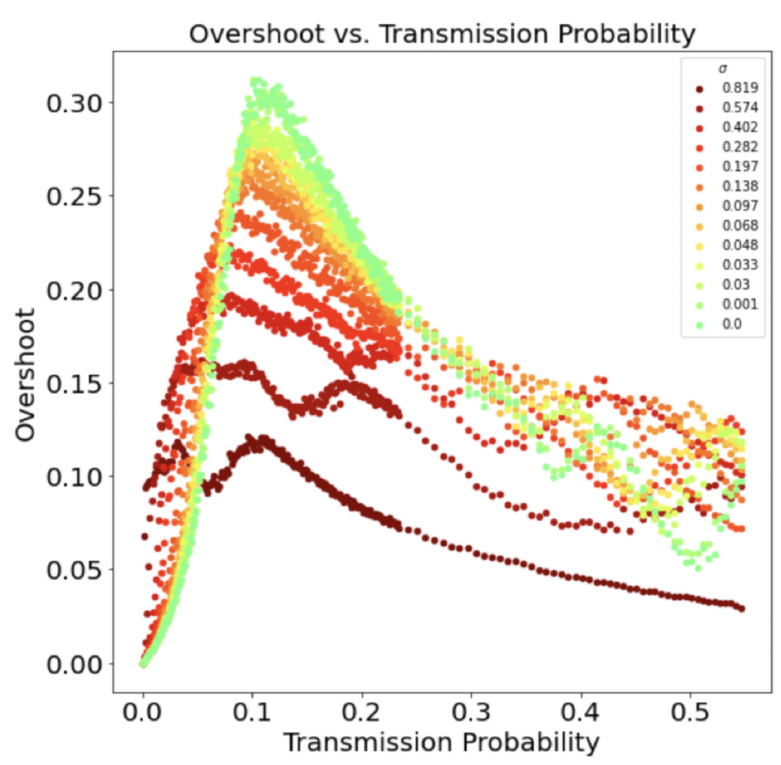
The overshoot for SIR epidemic simulations on networks with varying levels of heterogeneity (*σ*) as a function of transmission probability. *N* = 1000, *λ* = 5. Points represent the average of n = 150 simulation runs.

We also see that increasing contact heterogeneity qualitatively suppresses the overshoot peak both in terms of the overshoot value and the corresponding transmission probability. Furthermore, increased heterogeneity also flattens out the overshoot curve as a function of transmission probability.

## Discussion

We have proved that the maximum fraction of the population that can be infected during the overshoot phase of an epidemic in the Kermack-McKendrick SIR model is about 0.3, with a corresponding *R*_0_ ≈ 2.15. This upper bound on the overshoot seems to hold in other extensions of SIR models as well. In an SIR model with vaccinations, the upper bound stays the same. It also matches the numerical upper bound seen for SIR dynamics on networks of varying heterogeneity. In the 2-strain with vaccination SIR model of Zarnitsyna et. al. [17], the overshoot depends on both the level of strain dominance and vaccination rate, but from their results it is numerically seen that any amount of vaccination will produce an overshoot lower than the bound found here. Different control measures and strategies may reduce the overshoot as compared to the unmitigated case [7], keeping this upper bound intact. It will be interesting to see how general this bound is for SIR models with other types of complexities or for models beyond the SIR-type.

The mathematical intuition on why there is a peak in the overshoot as a function of *R*_0_ can be seen by an inspection of Equation 5. The first time term, 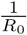 monotonically decreases with increasing *R*_0_. The last term, −*S*_*∞*_, monotonically increases with *R*_0_. Thus a trade-off in the two terms results in some intermediate peak. The epidemiological intuition behind a peak in the overshoot is that the total number of individuals infected during the epidemic grows monotonically with increasing *R*_0_. However, too high of an *R*_0_ leads to a sharp growth in the number of infected individuals, which burns through most of the population before the infection prevalence peak is reached, leaving few susceptible individuals left for the overshoot phase. This is seen by a monotonic decrease in the fraction of infected individuals that occur in the overshoot phase with increased *R*_0_ (Figure S2). Thus the maximal overshoot occurs as a trade-off between those two directions. From a public-health perspective, these results suggest that a significant number of infections may still occur after the peak of infections for diseases that spread relatively slowly (small *R*_0_).

## Data Availability

All data produced in the present work are contained in the manuscript.

## Methods

### Generating Graphs of Differing Heterogeneity

In Figure 6, we presented the results of SIR simulations of epidemics run on graphs of size N = 1000 and *λ* = 5, where the parameter of interest is *σ*. Each curve, from red to green, represents a different value of the graph heterogeneity. We implemented the following procedure from [12] for generating graphs as a function of a continuous parameter (*σ*):

The following simple procedure generates a graph that has the desired heterogeneity:

1. Choose values of *σ* (heterogeneity), *λ* (mean/median node degree), and *N* (number of nodes).
2. Draw N random samples from the following distribution using *σ* and *λ*, rounding these samples to the nearest integer, since the degree of a node can only take on integer values.

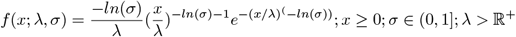
3. With the sampled degree distribution from the previous step, now use the configuration model method [11] (which samples over the space of all possible graphs corresponding to a particular degree distribution) to generate a corresponding graph.

This yields a valid graph with the desired amount of heterogeneity as specified by *σ*.

### Simulating Epidemics on Graphs of Differing Heterogeneity

We implemented the following simulation procedure from [12] for implementing an SIR epidemic on a graph: Given a graph *G*(*σ*) of heterogeneity *σ*, fix a transmission probability *τ* and recovery probability *γ*:

1. At time *t*_0_, fix a small fraction *f* of nodes to be chosen uniformly on the graph and assign them to the Infected state. The remaining (1 − *f*) fraction of nodes start as Susceptible.
2. For each *i* ∈ [1, *T*], for each pair of adjacent S and I nodes, the susceptible node becomes infected with probability *τ*.
3. For each *i* ∈ [1, *T*], each infected node recovers with probability *γ*.
4. At time *T*, record two quantities: The final attack rate and the herd immunity threshold at the peak of the epidemic.
5. Repeat steps (1-4) n = 150 times for each value of *τ*.
6. Repeat steps (1-5) for each value of *σ*.

### Numerical Solutions and Code

Numerical calculations and epidemic simulations on networks were performed in *MATLAB*. Code for all sections can be provided upon request.

## Acknowledgements

The authors would like to acknowledge Bryan Grenfell and Chadi Saad-Roy for their useful suggestions. The authors would like to acknowledge generous funding support provided by the National Science Foundation (CCF1917819 and CNS-2041952), the Army Research Office (W911NF-18-1-0325), and a gift from the William H. Miller III 2018 Trust.

## Author Contributions

M.M.N., A.S.F., S.A.O., S.A.L. designed research, performed research, and wrote and reviewed the manuscript.

## Additional information

The authors declare no competing interests.

## Supplemental Materials

### Supplemental Figures

**Figure S1.**
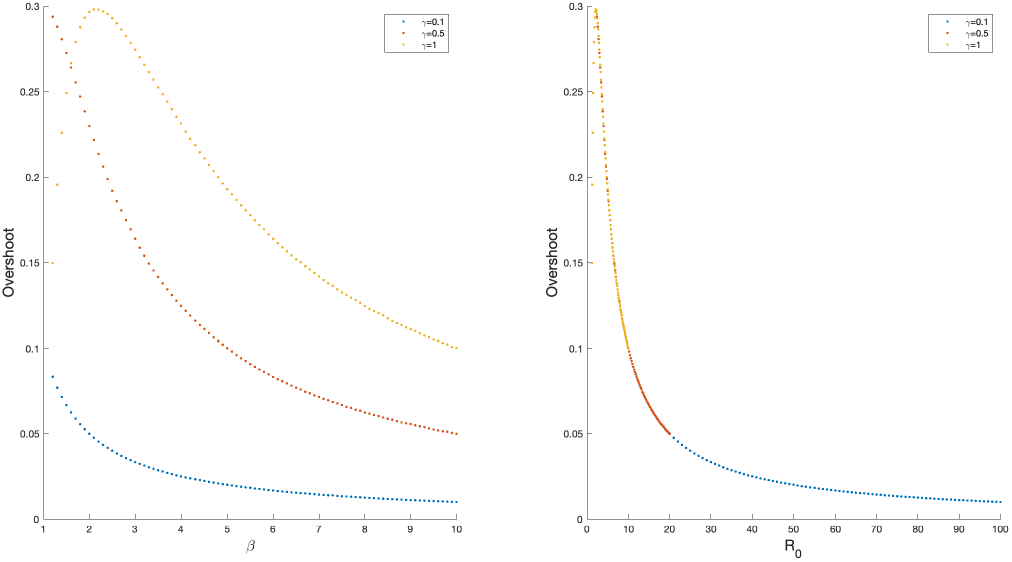
Comparing overshoot in the SIR model as a function of *R*_0_ compared to *β* and *γ* individually. Figure S1: a) The overshoot as a function of parameter *β* and different *γ*. b) The same data as in (a) but plotted in terms of 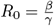.

**Figure S2.**
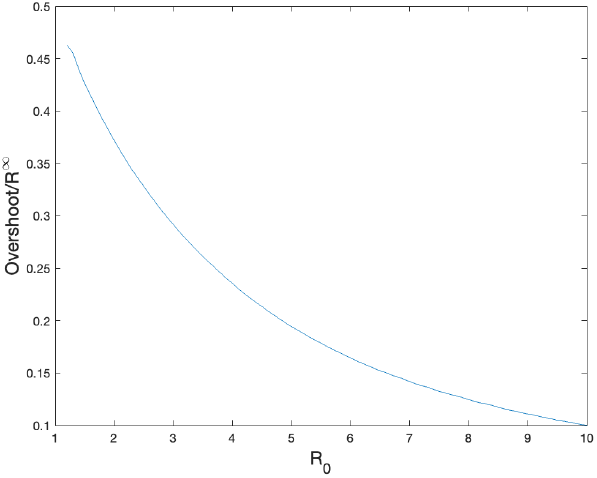
Fraction of individuals infected in the overshoot out of all individuals infected for SIR model. Figure S2: The ratio of overshoot to final attack rate as a function of *R*_0_.

